# Knowledge, attitudes, and practices of final-year healthcare students in low- and middle-income countries regarding epilepsy, dementia, and psychiatric disorders

**DOI:** 10.1101/2025.09.18.25336080

**Authors:** Emilie Auditeau, Marion Vergonjeanne, Leslie Cartz-Piver, Benjamin Calvet, Farid Boumédiène, Pierre-Marie Preux, KAPS Consortium

## Abstract

**Purpose:** Prevalence of chronic brain disorders, including dementia, epilepsy, and psychiatric disorders, is increasing worldwide. This represents a major challenge for future healthcare professionnals, especially in Low- and Middle-Income Countries (LMICs). This study aims to assess the knowledge, attitudes, and practices (KAP) of final-year students in medicine, pharmacy, and nursing specialties across three LMIC macro-regions to identify unmet needs and strengthen health education curricula.

**Methods:** A cross-sectional multicentric survey was conducted using a standardized, self-administered questionnaire developed by experts and translated into five languages (French, English, Spanish, Lao and Khmer). Administered through KoboToolBox, the questionnaire explored four domains (knowledge, attitudes, practices, and speciality) in dementia, epilepsy and psychiatric disorders. Scores are expressed as the average percentage of correct answers. Eligible participants were final-year students in medicine, pharmacy, and nursing.

**Results:** A total of 888 students from eight LMICs participated: 39.0% from Southeast Asia, 43.0% from Africa, and 18.0% from South America. Students’ mean age was 24.3 ± 4.7 years. The average percentage of correct answers was 76.1%, with significantly higher scores in psychiatric disorders (p < 0.001). Prior training and awareness were linked to higher KAP scores. Significant disparities were found by region, specialties, and disease, such as persistent misconceptions and stigma.

**Conclusion:** These findings will guide the incorporation of new teaching approaches in future healthcare education of dementia, epilepsy and psychiatric disorders in LMICs.

## Introduction

Chronic brain diseases, including epilepsy, dementia, and psychiatric disorders, affect 450 million people worldwide, causing substantial burden on individuals and society. In 2021, neurological disorders accounted for 11 million deaths and 443 million disability-adjusted life years (DALYs) [1]. This burden is pronounced in Low- and Middle-Income Countries (LMICs), where prevalence is high, and healthcare systems rely mainly on general practitioners, pharmacists, and nurses due to a shortage of specialized professionals.

Knowledge, Attitudes, and Practices (KAP) studies have assessed healthcare professionals’ preparedness for managing chronic brain diseases. Concerning epilepsy, multicenter studies in Brazil, United States, Portugal, Argentina and South Africa identified gaps in training and disease management, emphasizing a need for better education [2]. Studies in Saudi Arabia and Zambia demonstrated that higher education levels correlate with improved knowledge and reduced stigma [3,4]. Regarding dementia, KAP studies have focused on high-income countries, despite two-thirds of affected individuals living in LMICs [5]. Recent studies in South America, Asia, and Africa revealed knowledge gaps, particularly among pharmacy students, who are often overlooked in training programs [6–8]. For psychiatric disorders, stigma remains a major challenge, as shown in Nigerian studies where over 50% of individuals with depression concealed their illness for fear of discrimination [9]. Targeting final-year healthcare students aims at identifying gaps before entering the workforce and evaluating the effectiveness of current curricula.

## Objectives

The main objective of this study is to compare KAP scores of chronic brain diseases in LMICs. Secondary objectives are to (i) determine scores for knowledge, attitudes, practices and specialties by region (ii), to identify items with a low score and (iii) to determine factors associated with low scores.

## Methods

### Ethics statement

The study was authorized by the Ethics Committee of the Limoges University Hospital (number 398-2020-54) and by ethics committees of the eight participating countries.

### Study design

This is an observational, descriptive and cross-sectional study of healthcare students’ knowledge, attitudes and practices (KAP) at the end of their studies. The STROBE initiative for guidelines for observational studies in epidemiology (Strengthening the Reporting of Observational studies in Epidemiology, 2004) was the basis for developing and standardizing the study methodology.

### Setting

This study was conducted in eight LMICs among the network of the research laboratory EpiMaCT, Inserm U1094, IRD UMR270, Limoges University, France from 28 February 2020 to 13 March 2023. Volunteer LMICs included two South American countries (Peru, Ecuador), four African countries (Cameroon, Benin, Gabon, Madagascar) and two Southeast Asian countries (Cambodia, Laos). Most universities offered curricula in three healthcare specialties, and in some countries, public or private nursing schools were included. In each LMIC, a supervisor was identified among the University staff and a doctoral or postdoctoral student was designated to submit the KAP project to the local ethics committee for approval, identify and invite the eligible students and organize the on-site investigation.

### Participants

Eligibility criteria were the following: student enrolled in the selected universities and schools, in final year of medical, pharmacy or nursing studies. Healthcare students who had started an internship or had already obtained a diploma were not included. Student must have been physically present on the day of data collection and must have provided written informed consent after reading the information leaflet.

### Variables

The students filled out three questionnaires in the following order: epilepsy, dementia, and psychiatric disorders (**Supplements 1, 2, and 3**). Socio-demographic characteristics were collected, as well as data on previous training, awareness and contact with people living with the disease.

The questionnaire consisted of four domains: knowledge, attitudes, practices and specialization (KAP):

- Knowledge included definitions, epidemiological data and symptoms of the diseases,
- Attitudes included statements about social tolerance and stigmatization,
- Practices in diagnosis, management and medical treatment were included,
- Specialized items were designed for either medical, pharmacy or nurse students.

Each domain comprised five binary questions (Yes/No or True/False), with 1 point awarded for each correct answer and 0 for an incorrect one. The total score was calculated as the percentage of correct answers out of the 20 questions.

### Data sources/ measurement

The questionnaires were developed in French and pre-tested with a sample of midwifery students of University of Limoges. Versions in English, Spanish, Lao, and Khmer were created by forward-translation and back-translation. Validation of procedures took place in Gabon, in February to March 2020. The rest of data collection took place between September 2021 and July 2023. Data was collected online, using the KoboToolbox data collection tool. On the day of the investigation, a link was communicated to students to access the questionnaires online one by one. Each questionnaire was completed in an estimated time of 20 minutes. A short break took place between each. At the end of the work session, the investigators delivered an open lecture on epidemiological research concerning the three diseases, presenting examples of studies addressing disease knowledge, patient care pathways, and available treatments.

### Study size

This study used a convenience sampling method, with participant selection based on voluntary participation.

### Statistical methods

Checking and cleaning the generated database were performed after the export from KoboToolbox, using R. R and EasyMedStat (version 3.37.1) softwares were used for all statistical analyses and QGIS® was used for mapping. Counts and percentages were used to describe the categorical variables. For continuous quantitative variables, means and standard deviations were used. Chi² or Fisher exact tests were conducted to compare proportions. A two-factor ANOVA was conducted to compare participants’ scores across the three diseases based on sociodemographic variables, including gender, age, type of education, training, awareness, and experience of contact with an affected person. Levene’s test was done to compare variances and Shapiro-Wilk test to check the normality of continuous variables. For each disease, a multivariate linear regression was performed to assess the relation between the global score and the explanatory variables: age, gender, specialties, subcontinent, training, awareness, and contact with a person with the disease. Data were checked for multicollinearity with the Belsley-Kuh-Welsch technique. Heteroskedasticity and normality of residuals were assessed respectively by the White test and the Shapiro-Wilk test. The Newey West correction for heteroskedasticity was applied. A p-value < 0.05 was considered statistically significant.

## Results

### Participants

A total of 888 students living in the 8 LMICs participated, yielding 2348 answers. The study population included 346 students (39.0%) in Southeast Asia (SEA), 383 students (43.0%) from Africa (AFR) and 159 students (18.0%) from South America (SAM) (**Figure 1**). Among the eligible participants, participation level was 43.3% of SEA students, 47.7% of AFR students, and 66.0% of SAM students.

**Fig. 1.**
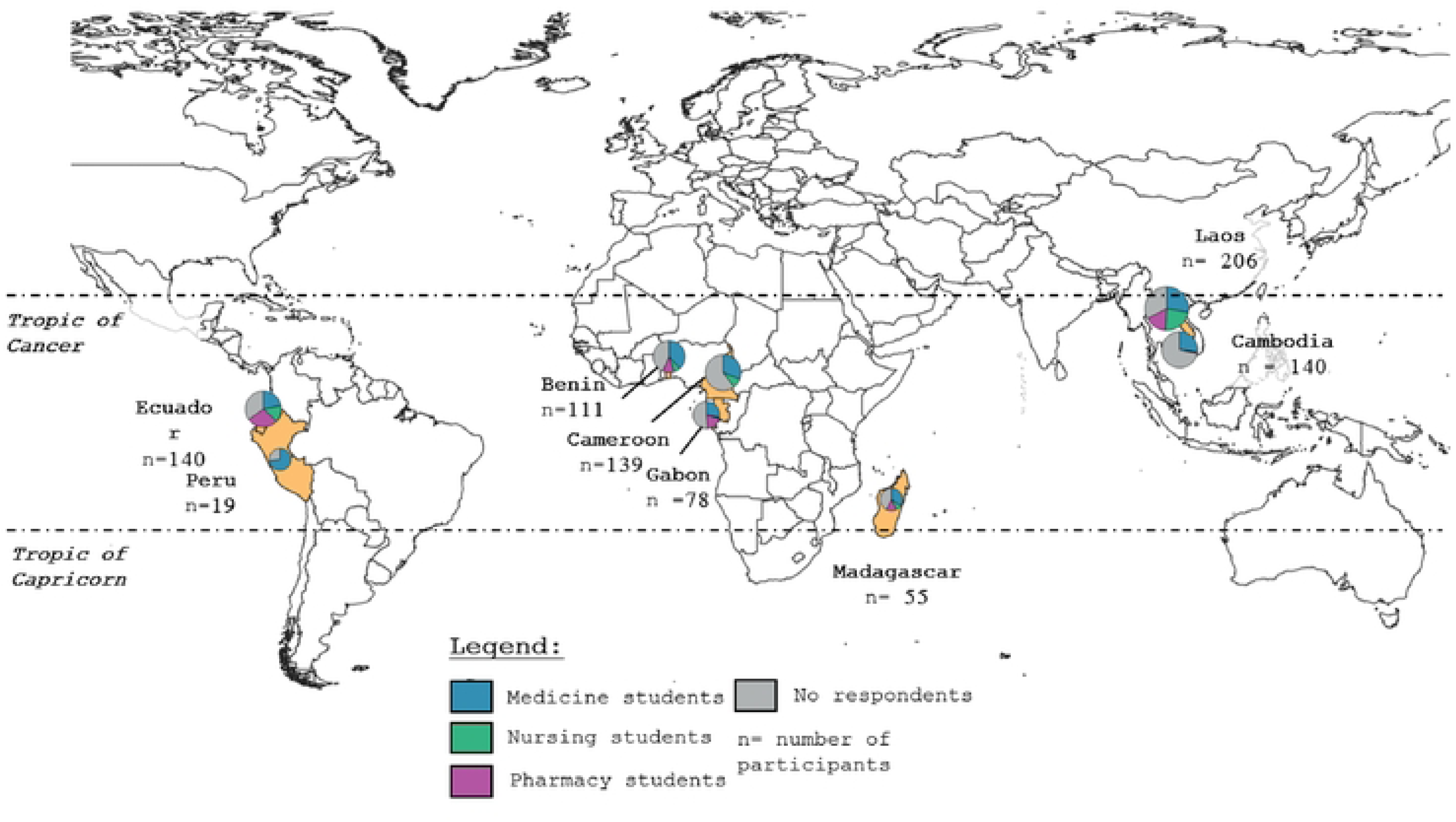
Study sites in South America, Africa and South East Asia.

### Socio-demographics, training, awareness and contact

Students’ average age was 24.3±4.7 years old, SAM students were significantly younger (p<0.001) (**Table 1**). Most students were female (60.5%). Medical students were the largest group with 530 students (61.1%), followed by nurses (20.9%) and pharmacy students (18.0%) (p<0.001). More SEA students reported training (epilepsy: 91%, dementia: 91.5%, psychiatric disorders: 89.5%) than students in SAM (78%; 77.3%; 73.4%) or in AFR (66.5%; 66.3%; 73.5%) (p<0.001). Dementia awareness was lower in AFR (83.2%) than in SAM (93.4%) and SEA (91.2%) (p=0.001). Awareness of psychiatric disorders was highest in SAM (99.3%) versus AFR (92.3%) and SEA (92.1%) (p=0.0024). Fewer students in SAM declared contact with a person living with epilepsy (44.0%) than in AFR (68.1%) or SEA (56.7%) (p<0.001). Contact with a person living with dementia was lowest in AFR (45.1%) versus SAM (45.3%) and SEA (54.2%) (p<0.001).

**Table 1.** Characteristics of the study population (n= 868)

| SOCIODEMOGRAPHIC DATA | AFRICA<br>(n=364) | SOUTH AMERICA<br>(n=159) | SOUTH EAST ASIA<br>(n=344) | TOTAL<br>(n=868) | P value |
| --- | --- | --- | --- | --- | --- |
|  | Mean ± SD<br>n (%) | Mean ± SD<br>n (%) | Mean ± SD<br>n (%) | Mean ± SD<br>n (%) |  |
| **Age (years)** | 24.5 ± 6.5 | 23.4 ± 2.1 | 24.5 ± 3.1 | 24.3 ± 4.7 |  |
| Less than 24 | 178 (48.9) | 103 (64.8) | 129 (37.5) | 410 (47.2) | <0.001<sup>a</sup> |
| More than 24 | 186 (51.1) | 56 (35.2) | 215 (62.5) | 458 (52.8) |  |
| **Specialties** |  |  |  |  | <0.001<sup>a</sup> |
| Medical | 247 (67.9) | 65 (40.9) | 218 (63.4) | 530 (61.1) |  |
| Nurses | 65 (17.9) | 62 (39.0) | 54 (15.7) | 181 (20.9) |  |
| Pharmacy | 52 (14.3) | 32 (20.1) | 72 (20.9) | 157 (18.0) |  |
| **Gender** |  |  |  |  | 0.366<sup>a</sup> |
| Female | 210 (57.7) | 103 (64.8) | 211 (61.3) | 525 (60.5) |  |
| Male | 154 (42.3) | 56 (35.2) | 133 (38.7) | 343 (39.5) |  |
| **EXPERIENCES WITH EPILEPSY** | n = 364<br>n (%) | n = 159<br>n (%) | n = 344<br>n (%) | n = 868<br>n (%) |  |
| **Training** |  |  |  |  | <0.001<sup>a</sup> |
| Yes | 242 (66.5) | 124 (78.0) | 313 (91.0) | 680 (78.3) |  |
| No | 122 (33.5) | 35 (22.0) | 31 (9.0) | 188 (21.7) |  |
| **Awareness** |  |  |  |  | 0.336<sup>b</sup> |
| Yes | 337 (92.6) | 153 (96.2) | 322 (93.6) | 813 (93.7) |  |
| No | 27 (7.4) | 6 (3.8) | 22 (6.4) | 55 (6.3) |  |
| **Contact with a person with the epilepsy** |  |  |  |  | <0.001<sup>a</sup> |
| Yes | 248 (68.1) | 70 (44.0) | 195 (56.7) | 514 (59.2) |  |
| No | 116 (31.9) | 89 (56.0) | 149 (43.3) | 354 (40.8) |  |
| **EXPERIENCES WITH DEMENTIA** | N= 315<br>n (%) | N= 137<br>n (%) | N= 284<br>n (%) | N= 736<br>n (%) |  |
| **Training** |  |  |  |  | <0.001<sup>a</sup> |
| Yes | 209 (66.3) | 106 (77.3) | 260 (91.5) | 575 (78.1) |  |
| No | 105 (33.7) | 31 (22.7) | 24 (8.5) | 160 (21.7) |  |
| **Awareness** |  |  |  |  | 0.001<sup>b</sup> |
| Yes | 262 (83.2) | 128 (93.4) | 259 (91.2) | 649 (88.2) |  |
| No | 53 (16.8) | 9 (6.6) | 25 (8.8) | 87 (11.8) |  |
| **Contact with a person living with dementia** |  |  |  |  | 0.0558<sup>a</sup> |
| Yes | 142 (45.1) | 62 (45.3) | 154 (54.2) | 358 (48.6) |  |
| No | 173 (54.9) | 75 (54.7) | 130 (45.8) | 378 (51.4) |  |
| **EXPERIENCES WITH PSYCHIATRIC DISORDERS** | N= 339<br>n (%) | N= 139<br>n (%) | N= 266<br>n (%) | N= 744<br>n (%) |  |
| **Training** |  |  |  |  | <0.001<sup>a</sup> |
| Yes | 249 (73.5) | 120 (73.4) | 238 (89.5) | 607 (81.6) |  |
| No | 90 (26.5) | 19 (26.6) | 28 (10.5) | 137 (18.4) |  |
| **Awareness** |  |  |  |  | 0.0024<sup>b</sup> |
| Yes | 313 (92.3) | 138 (99.3) | 245 (92.1) | 696 (93.5) |  |
| No | 26 (7.7) | 1 (0.7) | 21 (7.9) | 48 (6.5) |  |
| **Contact with a person living with a psychiatric disorder** |  |  |  |  | 0.1763<sup>a</sup> |
| Yes | 235 (69.3) | 85 (61.2) | 171 (64.3) | 491 (66.0) |  |
| No | 104 (30.7) | 54 (38.8) | 95 (35.7) | 253 (34.0) |  |

Legend: a) Chi<sup>2</sup> test; b) Fischer test

### Global KAP scores by disease: (**Table 2**)

Total scores showed 76.1% correct answers, with significantly higher scores for psychiatric disorders (p<0.001). Medical students performed better than pharmacy and nursing students (p<0.001). Regional differences were significant, with SEA students (73.0% ± 12.9) scoring lower than AFR students AFR (77.8% ± 11.0) and SAM students (78.5% ± 11.3) (p<0.001). Male students performed better in epilepsy and dementia (p=0.01). Training, awareness, and contact with patients significantly improved scores across all three diseases.

**Table 2.**
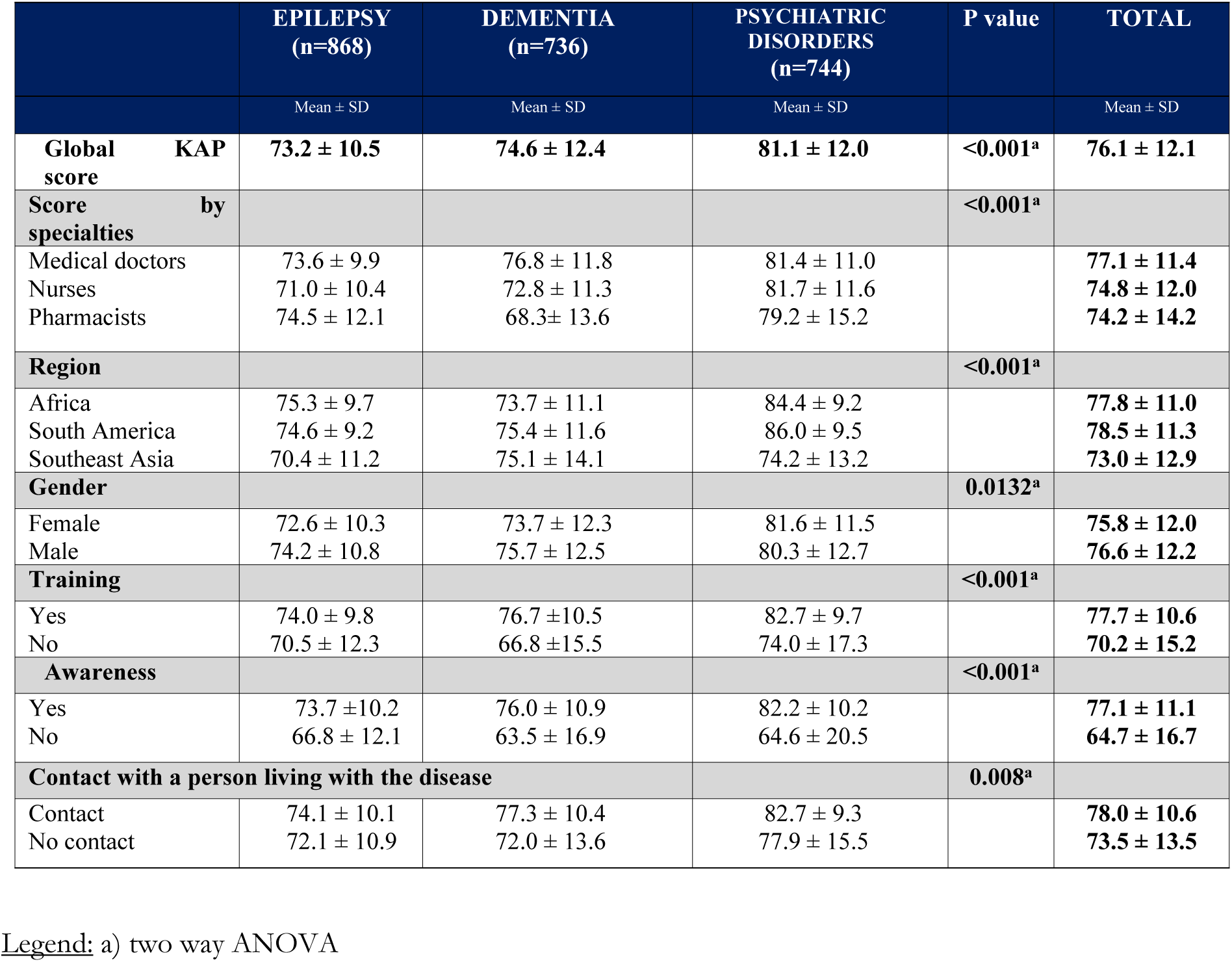
Mean percentages of correct answers by disease (epilepsy, dementia, psychiatric disorders)

|  | EPILEPSY<br>(n=868) | DEMENTIA<br>(n=736) | PSYCHIATRIC<br>DISORDERS<br>(n=744) | P value | TOTAL |
| --- | --- | --- | --- | --- | --- |
|  | Mean ± SD | Mean ± SD | Mean ± SD |  | Mean ± SD |
| **Global KAP score** | **73.2 ± 10.5** | **74.6 ± 12.4** | **81.1 ± 12.0** | **<0.001<sup>a</sup>** | **76.1 ± 12.1** |
| **Score by specialties** |  |  |  | **<0.001<sup>a</sup>** |  |
| Medical doctors | 73.6 ± 9.9 | 76.8 ± 11.8 | 81.4 ± 11.0 |  | 77.1 ± 11.4 |
| Nurses | 71.0 ± 10.4 | 72.8 ± 11.3 | 81.7 ± 11.6 |  | 74.8 ± 12.0 |
| Pharmacists | 74.5 ± 12.1 | 68.3 ± 13.6 | 79.2 ± 15.2 |  | 74.2 ± 14.2 |
| **Region** |  |  |  | **<0.001<sup>a</sup>** |  |
| Africa | 75.3 ± 9.7 | 73.7 ± 11.1 | 84.4 ± 9.2 |  | 77.8 ± 11.0 |
| South America | 74.6 ± 9.2 | 75.4 ± 11.6 | 86.0 ± 9.5 |  | 78.5 ± 11.3 |
| Southeast Asia | 70.4 ± 11.2 | 75.1 ± 14.1 | 74.2 ± 13.2 |  | 73.0 ± 12.9 |
| **Gender** |  |  |  | **0.0132<sup>a</sup>** |  |
| Female | 72.6 ± 10.3 | 73.7 ± 12.3 | 81.6 ± 11.5 |  | 75.8 ± 12.0 |
| Male | 74.2 ± 10.8 | 75.7 ± 12.5 | 80.3 ± 12.7 |  | 76.6 ± 12.2 |
| **Training** |  |  |  | **<0.001<sup>a</sup>** |  |
| Yes | 74.0 ± 9.8 | 76.7 ± 10.5 | 82.7 ± 9.7 |  | 77.7 ± 10.6 |
| No | 70.5 ± 12.3 | 66.8 ± 15.5 | 74.0 ± 17.3 |  | 70.2 ± 15.2 |
| **Awareness** |  |  |  | **<0.001<sup>a</sup>** |  |
| Yes | 73.7 ± 10.2 | 76.0 ± 10.9 | 82.2 ± 10.2 |  | 77.1 ± 11.1 |
| No | 66.8 ± 12.1 | 63.5 ± 16.9 | 64.6 ± 20.5 |  | 64.7 ± 16.7 |
| **Contact with a person living with the disease** |  |  |  | **0.008<sup>a</sup>** |  |
| Contact | 74.1 ± 10.1 | 77.3 ± 10.4 | 82.7 ± 9.3 |  | 78.0 ± 10.6 |
| No contact | 72.1 ± 10.9 | 72.0 ± 13.6 | 77.9 ± 15.5 |  | 73.5 ± 13.5 |

Legend: a) two way ANOVA

### Knowledge, Attitudes and Practices sections by disease

Concerning epilepsy, knowledge scores were lowest and similar among specialties (medical: 69.2%; pharmacy: 65.4%; nurses: 64.1%) whereas attitude and practice scores were good. Specialty scores were highest in pharmacy students (70.6%). (**Figure 2**) Concerning dementia, knowledge and attitude scores were good and comparable among specialties. Practice scores were lower in pharmacy (61.5%) than medical (77.0%) or nursing students (74.6%). Specialty scores were highest in medical students (82.3%) versus pharmacy (66.2%) and nursing (74.2%). For psychiatric disorders, scores were higher than for epilepsy or dementia. Knowledge and practice scores were excellent among medical students (K= 89.5%, A= 78.8%, P= 88.1%) and nurse students (K= 90.5%, A= 75.6%, P= 88.1%) and good in pharmacy students (K= 82.7%, A= 73.2%, P= 78.5%). Specialty scores were higher in pharmacy students (81.3%) than in medical (69.2%) or nursing students (72.2%). **Supplement 4**.

**Fig. 2.**
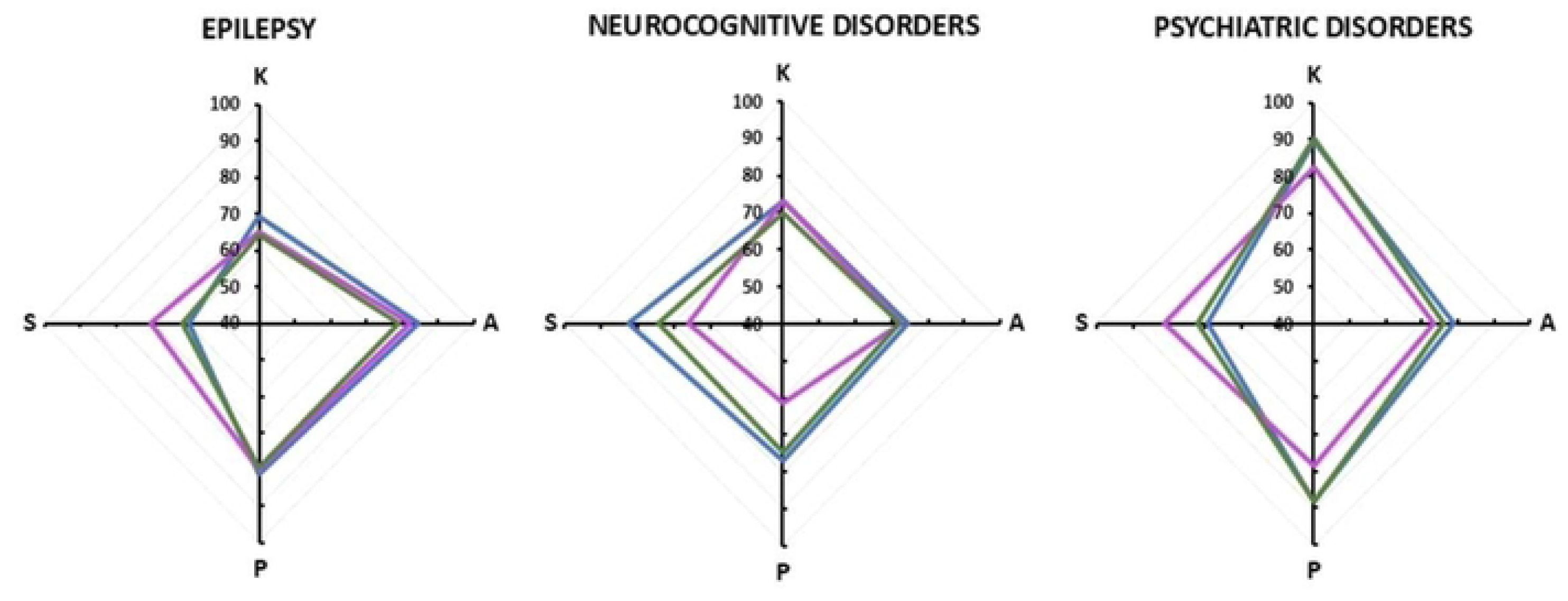
Percentages of correct answers by disorder and by specialty.

### Factors associated with a KAPS score

For the three diseases, previous training and awareness were associated with significantly higher KAP scores (**Table 3**). Training increased scores by 3.7% for epilepsy ([95% CI: 1.4; 5.9], p = 0.001), by 5.9% for dementia ([95% CI: 2.9; 8.8], p = 0.001), and by 8.7% for psychiatric disorders ([95% CI: 6.1; 11.3], p = 0.001). Awareness had an even stronger effect, increasing scores by 4.6% for epilepsy ([95% CI: 1.3; 7.8], p = 0.006), by 9.1% for dementia ([95% CI: 5.1; 13.1], p = 0.001), and by 10.8% for psychiatric disorders ([95% CI: 5.9; 15.6], p = 0.001).

**Table 3.**
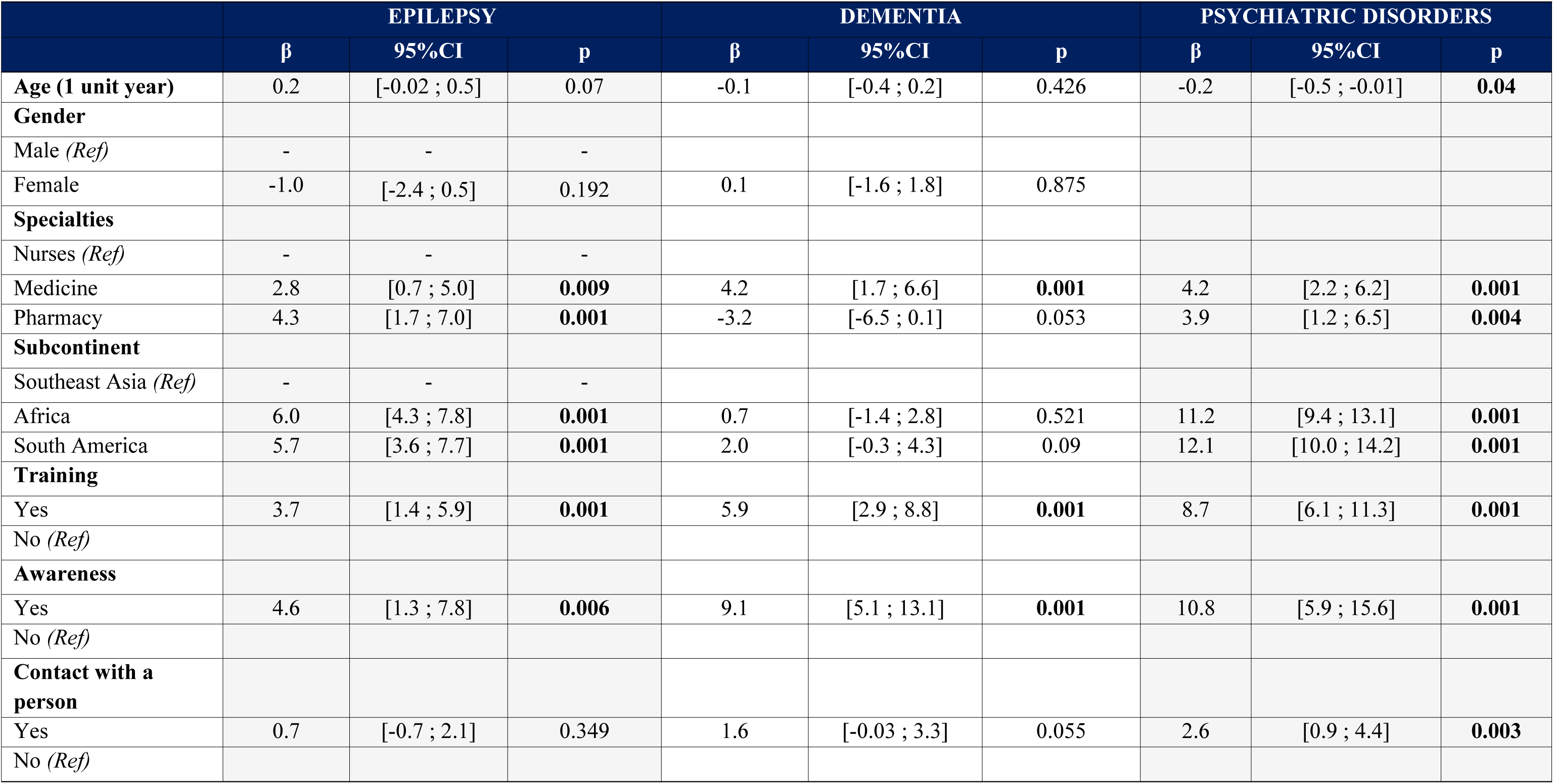
Factors associated with a good KAP score.

Only in epilepsy did the region influence KAP scores: SEA students scored significantly lower than AFR and SAM students. Finally, two factors were associated only to psychiatric disorders: contact with an affected person increased the KAP score by 2.6 points out of 100 ([95% CI: 0.9; 4.4], p = 0.003) and age had a significant negative impact, with each additional year reducing the KAP score by 0.2% ([95% CI: -0.5; -0.01], p = 0.04).

### Topics with low scores (less than 50% correct answers)

Low knowledge scores concerned disease definitions across all regions and all diseases (**Supplement 5**). Students struggled to define seizures and eclampsia in epilepsy, anxiety disorders in psychiatry, and questioned the curability of dementia in SEA. Attitude scores were lowest concerning contribution to society of individuals with epilepsy (AFR, SAM), dementia (AFR), and psychiatric disorders (AFR, SAM).

For epilepsy, practice scores were low in all regions regarding medication management. In psychiatric disorders, medication side effects scored low in AFR and SEA, where students also struggled with diagnosis. For dementia, students across all regions had trouble managing behavioral problems. Consideration of caregivers’ challenges showed low scores in AFR and SEA.

## Discussion

Our KAP study conducted among medical, pharmacy, and nursing students across three regions (Latin America, Africa, and Southeast Asia) revealed an overall satisfactory level of knowledge, attitudes, and practices regarding epilepsy, dementia and psychiatric disorders. The average global score was 76.1% (±12.1), with higher performance for psychiatric disorders (81.1 ± 12.0) compared to epilepsy (73.2 ± 10.5) and dementia (74.6 ± 12.4). Training and awareness significantly enhanced KAP scores for the three diseases, whereas contact with affected individuals improved scores for psychiatric disorders only.

Stigmatizing attitudes toward individuals with psychiatric disorders remain prevalent worldwide. This stigma can be linked to limited exposure to clinical psychiatry, insufficient training, and cultural biases. Tergesen et al. showed that exposure to video testimonials from individuals with psychiatric disorders significantly improved attitudes among medical students in Nepal [10]. Hanna et al. reported that structured educational programs led to more favorable perceptions among pharmacy students in the UK [11]. Zavorotnyy et al., demonstrated that structured psychiatric clerkships substantially improved medical students’ attitudes [12]. These educational strategies may also provide be valuable for addressing other stigmatizing conditions, such as epilepsy and dementia.

Regarding dementia, students in SEA demonstrated the lowest knowledge scores, indicating a need to strengthen theoretical understanding. Weakest areas included disease definition, symptom recognition, and awareness of the irreversible nature of dementia. A study conducted in Malaysia, including 464 medical students in final year, showed low overall dementia knowledge [13]. Higher knowledge of dementia was associated with previous formal dementia education (p=0.037) and experience in caring for dementia patients (p = 0.001). Our study revealed significant gaps in the management of behavioral disorders and caregiver support, with less than 50% correct answers. Similar deficiencies have been reported in previous studies; in Brazil, only 40.5% of students agreed to use depression medication for people living with dementia [19]. In previous KAP dementia studies, caregiver support was often omitted from the assessment tools, suggesting a need to integrate these components into educational curricula and evaluation frameworks.

Concerning epilepsy, knowledge remained the weakest domain (score = 66.2%), including definitions, seizure types, and first-line treatment. A study conducted in Lebanon among pharmacy students reported better results, with 93.1% students correctly identifying epilepsy and 78.7% demonstrating accurate knowledge of seizure characteristics [14].

### Strengths and limits

Voluntary participation of students may have led to selection bias, and the response rate of 48.2% among last year students could affect representativeness. Reliance on self-reported data introduces social desirability bias, which may have lead students to overestimate their knowledge or underreport negative attitudes. The cross-sectional design does not establish causality between education and KAP scores. Unlike longitudinal studies which track changes in students’ perceptions over time, our findings only provide a snapshot of students’ attitudes. The questionnaires were not formally validated even though pretesting was conducted. Due to COVID-19, data collection was prolonged and asynchronous across countries, potentially introducing temporal variability.

Despite these limitations, this study has several strengths. It includes final-year students from three specialities (medicine, pharmacy, nursing), providing a multidisciplinary perspective. The multicenter design across eight countries and three regions enhances the generalizability of findings. Investigating three stigmatizing diseases in a single study using standardized questionnaires translated into five languages ensures consistency and cultural adaptation. New themes, such as caregivers’ roles and patient inclusion, were introduced across the three diseases, as well as behavioral disorder management in dementia. Finally, the simultaneous exploration of knowledge, attitudes, and practices offers a comprehensive and comparative approach, setting this study apart from previous KAP research.

## Conclusion

This original KAP study highlights substantial disparities across different regions, healthcare specialties, and disease while also revealing several cross-cutting weaknesses: limited specialized training, insufficient clinical exposure, persistent misconceptions and enduring stigma. These findings underline the need to improve healthcare education by incorporating innovative pedagogical approaches such as patient simulations, testimonials and by increasing clinical exposure. Stigma-reduction initiatives should become a core component of healthcare professional training. Future research should focus on a longitudinal study design to assess the short and long-term impact of educational interventions on healthcare students’ knowledge, attitudes and practices, particularly in LMICs.

## Authors’ contributions

Conceptualization: EA, FB, PMP, LCP Data curation: EA, MV

Methodology/formal analysis/validation: FB, PMP, EA, MV

Project administration: FB, PMP, LCP

Funding acquisition: FB, EA

Writing – original draft: EA, MV, LCP

Writing – review & editing: FB, PMP, EA, MV, BC, LCP

## Conflict of interest

No potential conflict of interest relevant to this article was reported.

## Funding

This work was supported by Sanofi Accès aux Médicaments and Association ABIME. The funders had no role in study design, data collection and analysis, decision to publish, or preparation of the manuscript.

## Data availability

Dataset 1: Epilepsy questionnaire database Dataset 2: Dementia questionnaire database

Dataset 3: Psychiatric disorders questionnaire database

## Data Availability

All relevant data are within the manuscript and its Supporting Information files.

## Acknowledgments

We gratefully acknowledge the contributions of the Master’s students (M1 and M2) and the researchers who also participated in this project: Fousseyni Diadouba COULIBALY, Ludivine FAUGEROUX, Djouher HADJ-KADDOUR, Maria Bakita HOUEZE, Maryse HOUINATO, Euloge IBINGA, Argo PAMBUDI, Hélène SACCA and Ali SBEITY.

## Supplementary materials

Supplement 1: Questionnaire KAP Epilepsy (VE environ 9 pages)

Supplement 2: Questionnaire KAP Dementia (VE environ 9 pages)

Supplement 3: Questionnaire KAP Psychiatric Disorders (VE environ 9 pages)

Supplement 4: Mean percentages of correct answers by KAPS section and sub-continent

Supplement 5: Topics with low scores

## Notes

### Competing Interest Statement

The authors have declared no competing interest.

